# Clinical classifiers of COVID-19 infection from novel ultra-high-throughput proteomics

**DOI:** 10.1101/2020.04.27.20081810

**Authors:** Christoph B. Messner, Vadim Demichev, Daniel Wendisch, Laura Michalick, Matthew White, Anja Freiwald, Kathrin Textoris-Taube, Spyros I. Vernardis, Anna-Sophia Egger, Marco Kreidl, Daniela Ludwig, Christiane Kilian, Federica Agostini, Aleksej Zelezniak, Charlotte Thibeault, Moritz Pfeiffer, Stefan Hippenstiel, Andreas Hocke, Christof von Kalle, Archie Campbell, Caroline Hayward, David J. Porteous, Riccardo E. Marioni, Claudia Langenberg, Kathryn S. Lilley, Wolfgang M. Kuebler, Michael Mülleder, Christian Drosten, Martin Witzenrath, Florian Kurth, Leif Erik Sander, Markus Ralser

**Affiliations:** The Francis Crick Institute, Molecular Biology of Metabolism Laboratory, London NW11AT, United Kingdom; Department of Biochemistry, The University of Cambridge, Cambridge, CB21GA, United Kingdom; Charité Universitätsmedizin Berlin, Dept. of Infectious Diseases and Respiratory Medicine, 10117 Berlin, Germany; Charité Universitätsmedizin Berlin, Institute of Physiology, 10117 Berlin, Germany; Charité Universitätsmedizin Berlin, Core Facility - High Throughput Mass Spectrometry, 10117 Berlin, Germany; Charité Universitätsmedizin Berlin, Department of Biochemistry, 10117 Berlin, Germany; Department of Biology and Biological Engineering, Chalmers University of Technology, Gothenburg SE-412 96, Sweden; Berlin Institute of Health (BIH), and Charité Universitätsmedizin, Clinical Study Center (CSC), 10117 Berlin, Germany; Centre for Genomic and Experimental Medicine, Institute of Genetics and Molecular Medicine, University of Edinburgh EH4 2XU, United Kingdom; Usher Institute, University of Edinburgh, Nine, Edinburgh Bioquarter, 9 Little France Road, Edinburgh, EH16 4UX, United Kingdom; MRC Human Genetics Unit, Institute of Genetics and Molecular Medicine, University of Edinburgh, Edinburgh EH4 2XU, United Kingdom; MRC Epidemiology Unit, Institute of Metabolic Science, University of Cambridge, Cambridge CB2 0QQ, United Kingdom; Charité Universitätsmedizin Berlin, Department of Virology, 10117 Berlin, Germany; Department of Tropical Medicine, Bernhard Nocht Institute for Tropical Medicine, Hamburg, Germany

## Abstract

The COVID-19 pandemic is an unprecedented global challenge. Highly variable in its presentation, spread and clinical outcome, novel point-of-care diagnostic classifiers are urgently required. Here, we describe a set of COVID-19 clinical classifiers discovered using a newly designed low-cost high-throughput mass spectrometry-based platform. Introducing a new sample preparation pipeline coupled with short-gradient high-flow liquid chromatography and mass spectrometry, our methodology facilitates clinical implementation and increases sample throughput and quantification precision. Providing a rapid assessment of serum or plasma samples at scale, we report 27 biomarkers that distinguish mild and severe forms of COVID-19, of which some may have potential as therapeutic targets. These proteins highlight the role of complement factors, the coagulation system, inflammation modulators as well as pro-inflammatory signalling upstream and downstream of Interleukin 6. Application of novel methodologies hence transforms proteomics from a research tool into a rapid-response, clinically actionable technology adaptable to infectious outbreaks.

**Highlights:**

- A completely redesigned clinical proteomics platform increases throughput and precision while reducing costs.
- 27 biomarkers are differentially expressed between WHO severity grades for COVID-19.
- The study highlights potential therapeutic targets that include complement factors, the coagulation system, inflammation modulators as well as pro-inflammatory signalling both upstream and downstream of interleukin 6.

## Introduction

The ongoing SARS-CoV-2 pandemic has highlighted the pressing need for technologies that can accelerate our understanding of emerging diseases in order to (i) find markers that define disease severity, have prognostic value or define a specific phase of the disease, (ii) identify preventive strategies, and (iii) discover therapeutic targets. PCR-based diagnostics can be implemented and scaled quickly, but do not provide information about severity of the disease as well as likely illness trajectories (Chen et al., 2020; Corman et al., 2020). Furthermore, conventional biomarker as well as serological assays depend on affinity reagents such as antibodies. Developing these takes time and requires prior knowledge of epitopes and the disease mechanisms (Petherick, 2020). Mass spectrometry (MS)-based proteomics, instead, does not depend on affinity reagents and can be set up in an untargeted fashion, such that it does not depend on prior knowledge of the disease. It can quickly deliver substantial amounts of clinical and biological information from accessible biological material, such as blood plasma and serum. MS-based proteomics hence has the potential to become an ideal technology to be applied in situations when rapid responses are required. At present, MS-based proteomics workflows are well-established in research laboratories, where they are routinely used for biomarker discovery and profiling (Bruderer et al., 2019; Geyer et al., 2016a, 2016b, 2017, 2019; Liu et al., 2015; Niu et al., 2019; Wewer Albrechtsen et al., 2018). Increasingly, MS-based proteomics is also entering regulated clinical and diagnostic environments (Crutchfield et al., 2016). It has the potential to yield complex and predictive biomarker signatures that support clinical decision making, as well as to enable the prediction of patient trajectories via machine learning methods with datasets of sufficient depth and size (Ahadi et al., 2020). However, in clinical practice, its potential is yet to be completely realised. For routine application, MS-based proteomic methods must combine precision, reproducibility and robustness with low cost and high-throughput, such that results can be routinely compared within and between clinical studies and laboratories (Geyer et al., 2017, 2019; Nilsson et al., 2010; Wright and Van Eyk, 2017). These requirements might require a compromise with proteomic depth, which has often been a key objective of proteomics in research settings, but which has been, in turn, often achieved through long measurement times and high cost (Bruderer et al., 2019; Geyer et al., 2016a; Liu et al., 2015). A further hurdle is that current MS-based proteomic workflows require, partially due to their dependency on low flow-rate chromatography, a high level of expert knowledge to achieve the level of necessary robustness for implementation in the clinical laboratory.

Here we demonstrate a novel high-throughput mass spectrometry platform that enables quick and cost-effective in-depth analysis of disease susceptibility and progression in COVID-19 patients, as well as biomarker discovery. Our platform is redesigned at all steps from sample preparation, chromatography and data acquisition to data processing, in comparison to existing pipelines (Bache et al., 2018; Bekker-Jensen et al., 2020; Bian et al., 2020; Bluemlein and Ralser, 2011; Bruderer et al., 2019; Geyer et al., 2016a; Liu et al., 2015; Müller et al., 2020; Vowinckel et al., 2018). It uses a novel automated sample preparation workflow, that scales to high sample numbers through the use of liquid handling robotics and minimum hands-on-time, and includes effective strategies to mitigate longitudinal batch effects. Moreover, it renders short-gradient high-flow liquid chromatography (LC), a technology that is the basis of several FDA approved clinical assays (Grebe and Singh, 2011; Nair and Clarke, 2016), applicable to high-throughput proteomic experiments. Using this approach, we were able to reduce measurements to five-minute gradient length and used flow rates of 800μL/min, thereby substantially increasing sample turnover and reducing costs, while increasing stability and precision. We first benchmarked the platform on a cohort-based epidemiological study, Generation Scotland (GS) (Smith et al., 2013), and at considerable higher throughput, demonstrated a level of precision and consistency yet unachieved in large-scale proteomics. We then employed this new workflow in an immediate response to the SARS-CoV-2 pandemic outbreak in Germany, by applying it to a cohort which includes the first COVID-19 patients hospitalized at the Charité Universitätsmedizin, Berlin. We acquired measurements from a primary exploratory cohort comprising thirty-one COVID-19 patients studied to identify biomarkers (Figure 4a). We validated these on a smaller cohort of 17 independent patients and 15 healthy volunteers (Table S1). We identified biomarkers that would distinguish severe COVID-19 from milder forms of COVID-19 according to WHO grading, introduced as of April 2020 (World Health Organisation, 2020), and found several proteins, which have not been previously associated with COVID-19 severity. These are Alpha-1B-Glycoprotein (A1BG), Beta and Gamma-1 actin (ACTB;ACTG1), Monocyte Differentiation Antigen and Lipopolysaccharide co-receptor CD14, Lipopolysaccharide Binding Protein (LBP), Galectin 3 Binding Protein (LGALS3BP), Leucine-Rich Alpha-2-Glycoprotein (LRG1), Haptoglobin (HP), Protein Z-Dependent Protease Inhibitor (SERPINA10), Apolipoprotein C1 (APOC1), Gelsolin (GSN) and Transferrin (TF). Our results highlight the role of complement factors, the coagulation system, several inflammation modulators as well as pro-inflammatory signalling both upstream and downstream of interleukin 6, and identify new prognostic biomarkers for COVID-19. In addition, our findings validate a subset of protein biomarkers that were reported in a parallel study for a cohort recruited at an earlier stage of the pandemic in China, analysed with a conventional and more time consuming proteomics technology (Shen et al., 2020), as well as those found to correlate with COVID-19 with conventional serological investigations (Chen et al., 2020; Shi et al., 2020; Zhou et al., 2020a). Our study demonstrates the value and power of robust high-throughput mass spectrometry in a global public health crisis. Very fast and reliable proteome technologies can play a vital role both in clinical classification, as well as in the rapid identification of new therapeutic targets against novel infecting agents.

## Results and Discussion

### 1. A new platform for high-throughput large-scale proteomics

We addressed throughput, precision, costs and practical hurdles in clinical implementation of MS-based proteomics, by designing a new proteomics platform, redesigning sample preparation, chromatography, mass spectrometric acquisition and data analysis. Our workflow starts after the transfer of the clinical plasma/serum samples, obtained with standard operating procedures, to 96-well plates, upon which all pipetting and mixing steps are conducted with liquid handling robots. The preparation workflow builds on a simple but effective innovation that mitigates batch variation in sample preparation reagents, which has proven so far a major and prohibitive contributor to quantification inconsistencies in (large-scale) proteomic experiments (Fu et al., 2018; Lowenthal et al., 2014; Piehowski et al., 2013) (Figure 1). Instead of pipetting new reagents on the samples at each step, initial common stock solutions (urea/ammonium bicarbonate buffer/dithiothreitol, iodoacetamide, formic acid and trypsin) are pre-filled into multiwell plates and are then stored at −80°C for whole projects in perpetuity. These plates enter the workflow at different stages (Figure 1), thereby not only reducing hands-on time, but maintaining the exact same reagents for projects of, in theory, unlimited scale. A second novelty is that clean-up of the digested peptides is done with 96-well solid-phase extraction plates, of which four are processed in parallel to reduce technical variability. Finally, the inclusion of sample preparation controls on each plate enables cross-batch normalisation, to correct batch-effects in case these emerge at the acquisition step of the platform (Figure 1).

The next challenge was to establish a data acquisition scheme that could be implemented in regulated environments without major hurdles and maintain very low variability across large sample series. Here, we focused on the implementation of a new chromatographic regime. The current standard technique for bottom-up proteomics, nano-flow liquid chromatography, used owing to its high sensitivity, is a main contributor to batch variability in liquid chromatography-mass spectrometry (LC-MS) experiments (Gama et al., 2013; Shishkova et al., 2016). Several recent studies have shown that run-to-run variability improves by switching from nano-flow to micro-flow regimes, or to specialized chromatographic devices that operate with pre-formed gradients. These allow faster runtimes and sample turnover, show better retention time stability, and improve column lifetime (Bache et al., 2018; Bian et al., 2020; Bruderer et al., 2019; Vowinckel et al., 2018). It came to our attention that high-flow LC (also known as analytical LC) in conjunction with very fast chromatographic gradients, a technology that reaches the requirements of regulated clinical laboratories, could further and substantially improve throughput and chromatographic properties. However, typically it has not been applied to short-gradient proteomics for a number of reasons. Firstly, on this type of fast chromatography, conventional mass spectrometric acquisition schemes do not reach sufficient sampling velocity in data-dependent mode (peaks elute too fast). Secondly, when using data-independent acquisition schemes which do not sample each peak individually, conventional software cannot deconvolute the interference-rich short-gradient data produced (Demichev et al., 2020; Messner et al., 2019). We were able to overcome these issues, and present an acquisition scheme that bases on 5-minute water to acetonitrile chromatographic gradients at a flow rate of 800 μl/min. Separating tryptic digests of non-depleted human plasma using an 1290 Infinity II UPLC system (Agilent) coupled to a TripleTOF 6600 (Sciex) mass spectrometer, illustrates that Total Ion Chromatograms (TICs) are virtually unchanged over repeated injections (Figure 2a). The nearly complete overlap of the TICs indicates not only the stability of the applied chromatographic system, but also the stability of the electrospray, that is facilitated by the gases that assist the desolvation process in high-flow ion-sources. By using a short 5 cm column and by increasing the flow rate post-gradient to 1200 μl/min and 1000 μl/min during washing and equilibration, respectively, we were able to reduce the total runtime, including overheads, to < 8 minutes. In this particular test, the setup allows a theoretical throughput of 180 samples/day on a single mass spectrometer, an at least five-fold improvement compared to microLC or nanoLC platforms optimized for throughput (Bruderer et al., 2019; Geyer et al., 2016a; Vowinckel et al., 2018). Moreover, the columns used in high-flow chromatography have higher capacities and thus are less prone to carryover. Indeed, blank injection after 10 acquisitions of non-depleted plasma tryptic digests, shows no significant carryover even with an applied wash time of less than one minute (Figure 2a).

The separation of a K562 cell line tryptic digest (Promega) at different gradient lengths illustrates the chromatographic properties achieved. The high-flow setup used achieved a median peak Full Width at Half Maximum (FWHM) of 3 seconds with a 20-minute gradient length. For comparison, an extensively optimized micro-flow LC (Demichev et al., 2020; Messner et al., 2019), achieved a FWHM of 5 seconds, at the same gradient length (Figure 2c). Thus, high-flow gradients as fast as 5 minutes resulted in peak capacities comparable to the highly optimized 20-minute micro-flow setup (Demichev et al., 2020; Messner et al., 2019) (Figure 2d).

In order to achieve a sufficiently fast mass spectrometric duty cycle, we used a q-TOF instrument with a very fast sampling rate (Schilling et al., 2017), and applied SWATH-MS, a data-independent acquisition (DIA) method specifically developed to minimize stochastic elements in data acquisition (Gillet et al., 2012; Ludwig et al., 2018). In order to record sufficient data points per chromatographic peak, we optimized the method for duty cycles of 700 milliseconds, and scan a precursor mass range of m/z 450–850 using 25 windows with variable window size and with 25 ms accumulation time. In order to deconvolute the complex data recorded, we built on our recent developments of DIA-NN software, that includes several algorithms that boost the number of true positive precursor identifications in the short-gradient DIA-MS runs. DIA-NN can handle complex short gradient data as it contains novel algorithms that correct for signal interferences, and uses Deep Neural Networks to assign the confidence scores to peptide-spectrum matches and are able to correctly classify true positive signals (Demichev et al., 2020). In order to further improve DIA-NN (version 1.7.10) for the high-throughput workflow presented here, we have implemented the MaxLFQ protein quantification algorithm (Cox et al., 2014). Originally, it was designed for shotgun-MS studies, but was recently introduced to DIA proteomics (Pham et al., 2020) and increased the quantification precision for serum and plasma proteomes.

**Figure 1:**
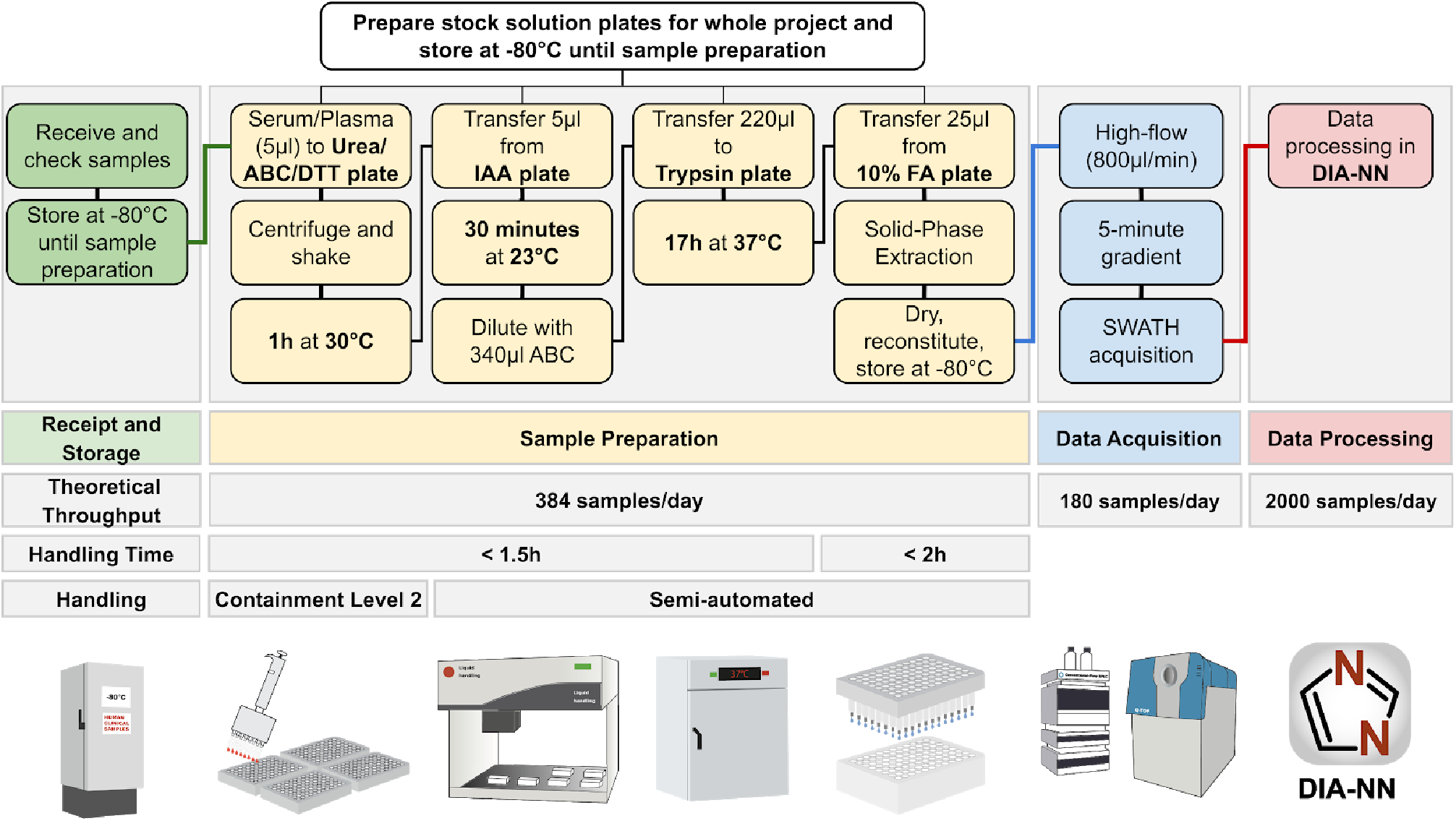
A redesigned high-throughput proteomics platform for large-scale and longitudinal clinical proteomic studies. **Receipt and Storage (green boxes):** Clinical or Epidemiological samples are collected using a standard operating procedure, received and stored at −80°C, then aliquoted to 96-well plates alongside control samples. For plasma and serum, 5μL are processed and yield sufficient tryptic digest for five measurements on the high-flow rate LC-MS platform. **Sample Preparation (yellow boxes):** The sample preparation workflow is designed for handling 384 samples/batch (four 96-well plates). Batch effects are mitigated by using pre-aliquoted stock solution plates - prepared for whole projects and stored at −80°C - that enter the workflow at different steps, as well as by using a liquid handling robot for pipetting and mixing. Sample clean-up is done with 384 samples/batch by using 96-well solid-phase extraction plates (BioPureSPE, The Nest Group) and a liquid handler for pipetting. The hands-on time for clean-up is < 2 hours and although the digestion is done overnight, the total hands-on time for the sample preparation is < 3.5 hours. **Data Acquisition (blue boxes):** ultra-fast measurements of the digested samples are facilitated in 300 seconds chromatographic gradients using high-flow chromatography (800μl/min) with a short reversed phase C18 column (50mm × 2.1mm, 1.8 μm particle size) to accelerate equilibration and washing steps. A 700 milliseconds duty cycle, required to record sufficient data points per chromatographic peaks, that elute at full width at half maximum (FWHM) of about 3 seconds, is achieved with an optimized SWATH data acquisition method. The theoretical throughput of data acquisition for one mass spectrometer is 180 samples/day. **Data Processing (red boxes):** The analysis of the highly complex short-gradient DIA data is achieved with an optimized version (1.7.10) of DIA-NN (Demichev et al., 2020). DIA-NN is based on neural networks to enable confident peptide identification with fast gradients and achieves a throughput of > 2000 samples/day on a conventional PC. Abbreviations: ABC, ammonium bicarbonate; DTT, dithiothreitol; IAA, iodoacetamide; FA, formic acid.

**Figure 2:**
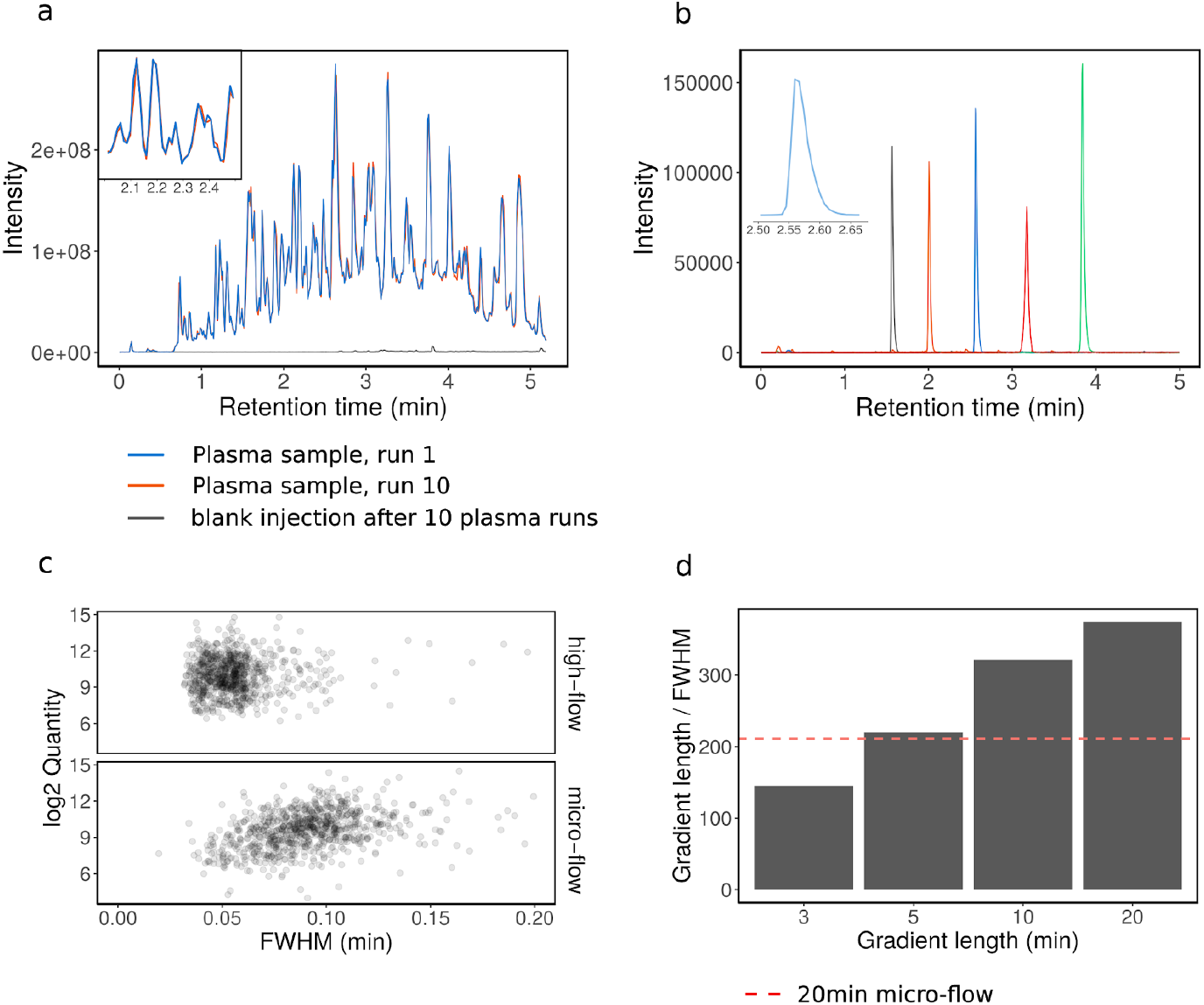
High-flow liquid chromatography (LC) and its application to short-gradient MS based proteomics. **a**. Tryptic digests of 10 human blood plasma samples were injected and separated with a 300 seconds linear water to acetonitrile chromatography gradient using an Agilent 1290 Infinity II LC system coupled to a TripleTOF 6600 mass analyzer. The total ion chromatograms (TIC) of the first and last injection were overlaid and coloured with blue and red, respectively. The time from the start of one run to the next was reduced to 8 minutes (including instrument overheads), which enables a throughput of ~180 samples/day. After the 10 plasma injections, water was injected and the TIC (black line) shows no significant carryover despite the short washing time. **b**. Extracted ion chromatograms of 5 synthetic peptides (AETSELHTSLK (m/z 408.55, black line), LDSTSIPVAK (m/z 519.80, orange line), ALENDIGVPSDATVK (m/z 768.90, blue line), AVYFYAPQIPLYANK (m/z = 883.47, green line) and TVESLFPEEAETPGSAVR (m/z 964.97741, red line) from a synthetic peptide mixture (Pepcal, Sciex) as separated on the 300 seconds linear gradient. Chromatograms were extracted from TOF MS data, width = 0.1 Da. **c**. A tryptic digest of K562 human cell lines was separated with a 20-minute linear gradient ramping from 3% ACN 0.1% FA to 36% ACN, 0.1% FA on high-flow (800 μl/min; C18 column 50mm × 2.1, column length). Peak widths at full width at half maximum (FWHM) of the eluting peptides were compared to a 20-minute micro-flow run (5 μl/min; 15 cm column (Demichev et al., 2020)), analysed on the same mass spectrometer (Sciex TripleTOF 6600). **d**. Peak capacities (Gradient length / FWHM) for 3, 5, 10 and 20-minute linear gradients (3% ACN/0.1% FA to 36% ACN/0.1% FA) on high-flow compared to 20-minute micro-flow chromatographic gradients (red dashed line).

### 2. Benchmarking acquisition depth, quality and data consistency in an epidemiological study

To assess the suitability of our high-throughput platform for human blood plasma and serum proteomics, we generated proteomes for undepleted serum samples derived from 199 random individuals that participated in the Generation Scotland (GS) epidemiological study (Smith et al., 2013). GS is a family-based cohort of approximately 24,000 individuals in 7,000 family groups from across Scotland, aged between 18 and 98 (Smith et al., 2013). We also included a large number of commercial plasma (tebu-bio, 91 total) and serum (tebu-bio, 79 total) samples as quality controls for the sample preparation workflow, as well as repeated injections of a single sample every 11 samples (pooled from 32 prepared commercial serum samples, 39 total) as a control for the LC-MS performance. The sample preparation was done in four 96-well plates and the experiment involved 409 non-blank injections.

After processing the raw data with DIA-NN using a high-quality spectral library (Bruderer et al., 2019), we assessed the robustness and consistency of protein identification achieved. The high-flow LC setup yielded exceptional retention time stability across the whole experiment (Figure 3a). As expected, due to the short gradients, the total proteomic depth (total number of peptides quantified) is lower than achieved with MS workflows that use pre-fractionation and longer gradients with lower flow rates, and have a slower duty cycle to scan over a larger mass range. In undepleted plasma, these typically detect 250 to 450 proteins per injection (Bian et al., 2020; Bruderer et al., 2019; Geyer et al., 2016a; Liu et al., 2015). However, with an average of ~270 protein groups detected per injection and 311 in total, our much faster platform still covers most of the typical plasma proteome also seen by the other methods, and quantifies about 50 FDA approved biomarkers (Anderson and Anderson, 2002). Moreover, for large-scale experiments, the numbers of consistently quantified peptides and proteins are more relevant than the maximum number of protein groups identified, as only consistent detection allows for quantitative comparison between individuals and is suitable for the development of clinical assays. In these measures, the platform performs exceptionally well. We consistently identified around 3,000 peptide precursors (i.e. peptides ionised to a specific charge; Figure 3b) and 200 unique proteins (i.e. gene products identified with specific proteotypic peptides; Figure 3c) across all 409 proteome acquisitions. In total we detected 311 protein groups, out of which 245 uniquely identified proteins were measured with 87% data completeness. Among these, 182 unique proteins were measured with 99% data completeness (Figure 3d).

**Figure 3.**
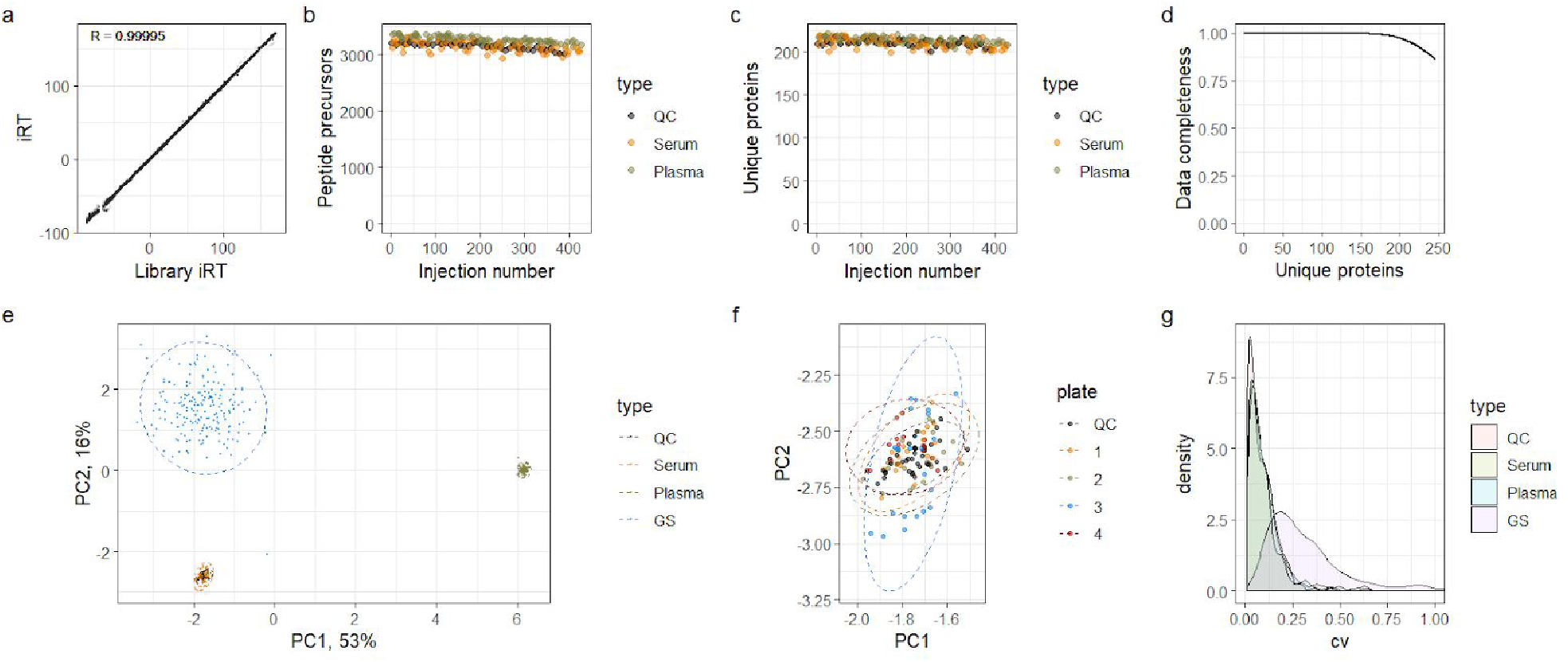
Robustness and quantitative precision of a new clinical proteomic platform applied to a population-based epidemiological cohort. 409 serum proteomes were analysed for characterizing 199 participants of the GS study. The sample series is composed of 39 repeat injections (“QC”), 79 serum and 91 plasma commercial sample preparation controls and 200 patient serum samples derived from the 199 individuals (“GS”). **a**. Overlayed aligned retention times (Biognosys iRT scale) of all peptide identifications in the whole experiment. Median iRT standard deviation (SD) was 0.22 (relative SD = 0.0009) and correlation between the observed iRT and library iRT was 0.99995, indicating very high retention time stability. **b**. Numbers of peptide precursors and **c**. unique proteins identified in control samples. **d**. Data completeness in the whole experiment plotted against the number of proteins identified. The data completeness for all 245 unique proteins was 87%, while 182 proteins were identified with data completeness 99%. **e**. PCA using consistently identified proteins (log-transformed quantities). **f**. The “Serum” cluster on the PCA plot, with samples prepared on different 96-well plates coloured differently. No bias between the plates can be detected. **g**. Coefficient of variation (CV). After accounting for instrument drift, median CV values are 5.4% for replicate injections (“QC”), 7.6% for serum controls, 7.3% for plasma controls and 25.6% for the participants’ samples.

To assess the quantitative precision, we first illustrated the proteomic data using principal component analysis (PCA, Figure 3e). The PCA data fully separates in PC1 all serum from plasma samples, and in PC2, the control serum samples from the GS serum samples. The difference between the GS samples and the commercial samples might be explained by different serum collection/storage procedures. Moreover, the biological variability across the randomly chosen individuals is much higher than the technical variability (spread of GS samples vs Serum/Plasma samples), and hence is detected with high confidence by our platform. We further examined in detail the “Serum” cluster of points, and did not detect any bias between different sample preparation plates (Figure 3f).

Finally, we evaluated the quantification precision by calculating the coefficient of variation (CV) of protein quantities across the studies. Median values obtained are 5.4% for repeat reference sample injections (“QC”) after instrument drift correction, 7.6% for serum controls and 7.3% for plasma controls, reflecting the precision of the entire workflow including sample preparation, acquisition, and data analysis. These values are much lower than the biological variation detected across the randomly chosen GS participants, which show a CV of 25.6% (Figure 3g). The platform hence confidently identifies biological variability in large-scale serum proteomic experiments of randomly chosen and presumed healthy individuals. Indeed, to our knowledge, such high precision values (5.4% CV for the LC-MS part of the workflow, 7.3% for the entire workflow including sample preparation over processing 409 proteomes) have not been achieved to date in comparable large-scale proteomic studies.

### 3. Rapid and precise high-flow rate proteomics identifies novel biomarkers for COVID-19

We applied the new workflow for the analysis of serum and citrate plasma samples for two independent COVID-19 cohorts that included patients who were among the first that were hospitalized at Charité – Universitätsmedizin Berlin, between March 1st and March 26th 2020. Thirty-one SARS-CoV-2 infected patients were included in the exploratory cohort to identify biomarkers (Figure 4a). 11/31 (35 %) patients were female and 20/31 (65 %) were male, median age was 54 years (range 21–81). Severity of COVID-19 was graded using the WHO ordinal outcome scale of clinical improvement (score 3 = hospitalised, no oxygen therapy, score 4 = oxygen by mask or nasal prongs, score 5 = non-invasive ventilation or high-flow oxygen, score 6 = intubation and mechanical ventilation, score 7 = ventilation and additional organ support - pressors, RRT, ECMO) (World Health Organisation, 2020). (Table S1 for cohort, Table S2 for WHO criteria). Four patients (13 %) died from COVID-19, and 4 patients remain hospitalized at the time of writing. All other patients have been discharged in good health from hospital. Successively, a control group, consisting of 15 healthy volunteers, and 17 further patients suffering from COVID-19, was recruited at the same hospital, and used for validation of the biomarkers discovered (Table S1).

**Figure 4.**
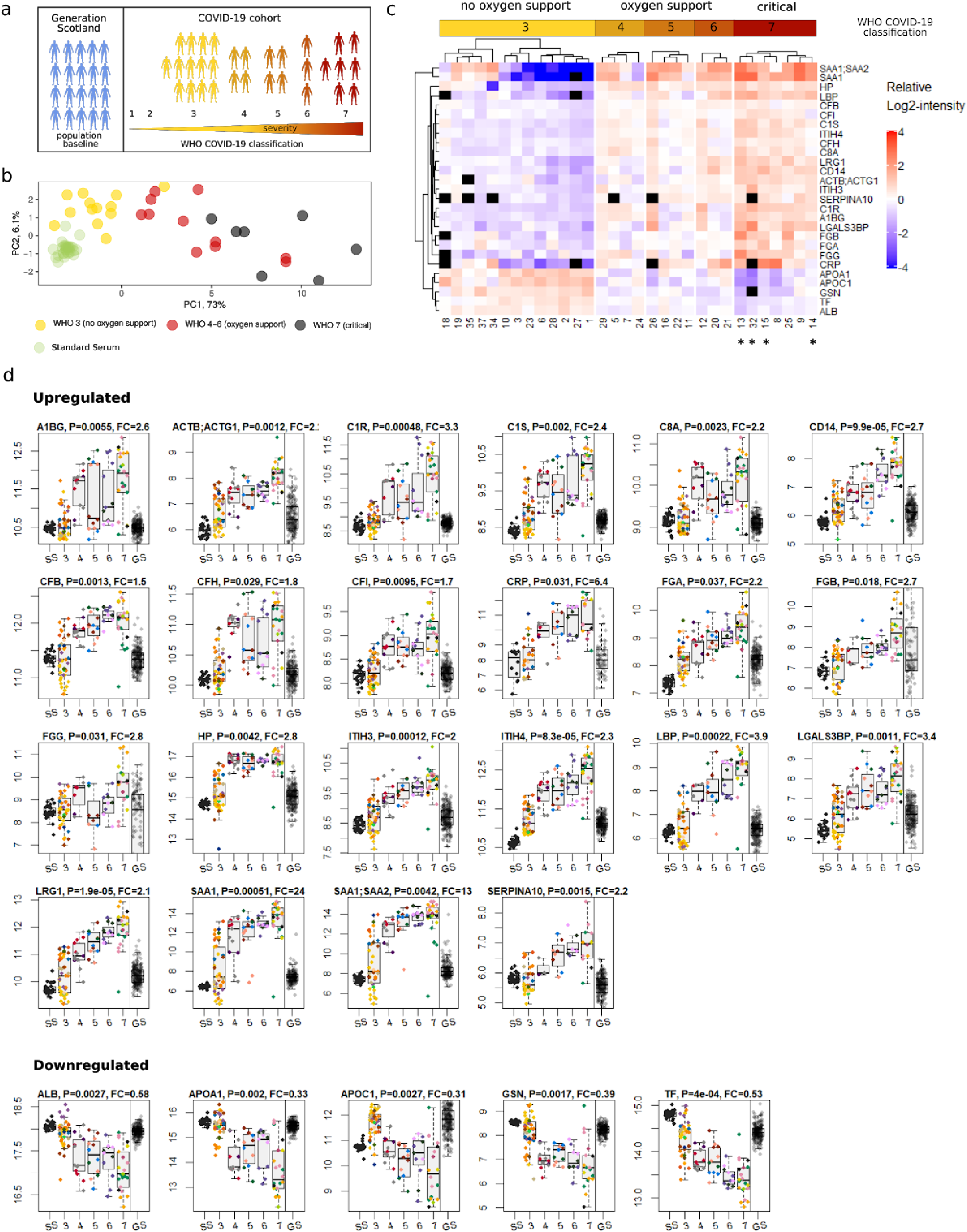
Protein biomarker signatures of COVID-19 severity. **a**. Study design. 199 random individuals from the Generation Scotland (GS) study were measured to assess the performance of the platform and to obtain a population baseline. Protein responses based on COVID-19 severity were obtained from a cohort of 31 hospitalized SARS-CoV-2 infected patients. Severity of COVID-19 was graded using the WHO ordinal outcome scale of clinical improvement (World Health Organisation, 2020). **b** Principal component analysis (PCA) based on genes found differentially expressed depending on COVID-19 severity. Median quantities across all time points were calculated for each patient and 29 genes without missing values were used to generate the PCA plot (quantities were standardized). Cases with the severity “3” on the WHO scale (hospitalized, no oxygen therapy) are well separated from cases with the severity “7” along the first principle component, with “4” - “6” cases in-between. **c** Heatmap shows protein signatures that report on COVID-19 severity. Visualisation was performed using the ComplexHeatmap R package (Gu et al., 2016). Black‘squares’ indicate missing values. Patients labelled with an asterisk (^*^) had a fatal outcome of the disease. **d**. Proteins upregulated (top panel) and downregulated (lower panel) depending on COVID-19 severity (WHO grade; SS - standard serum: GS - Generation Scotland), as well as the population spread of the protein abundance in 199 randomly selected individuals of an independent cohort (Generation Scotland; GS). As the absolute quantities from the COVID-19 and GS studies cannot be compared directly (samples were obtained in a different manner), we simplified the visual assessment of the population spread, by normalizing by the median of GS quantities to the median of WHO grade 3 (no oxygen support) COVID-19 cases (the normalized values were used for illustration purposes only, and not used for testing for statistical significance). **Proteins Upregulated with increasing severity of COVID-19:** A1BG (Alpha-1B-Glycoprotein), ACTB;ACTG1 (Actin Beta and Gamma-1), C1R (Complement C1r), C1S (Complement C1s), C8A (Complement C8 Alpha Chain), CD14 (Monocyte Differentiation Antigen CD14), CFB (Complement Factor B), CFH (Complement Factor H), CFI (Complement Factor I), CRP (C-Reactive Protein), FGA, FGB and FGG (Fibrinogen Alpha, Beta and Gamma Chains), HP (Haptoglobin), ITIH3 (Inter-Alpha-Trypsin Inhibitor Heavy Chain 3), ITIH4 (Inter-Alpha-Trypsin Inhibitor Heavy Chain 4), LBP (Lipopolysaccharide Binding Protein), LGALS3BP (Galectin 3 Binding Protein), LRG1 (Leucine-Rich Alpha-2-Glycoprotein), SAA1 (Serum Amyloid A1), SAA1;SAA2 (Serum Amyloid A1 and A2 protein group), SERPINA10 (Protein Z-Dependent Protease Inhibitor); **Proteins Downregulated with increasing severity of COVID-19:** ALB (Albumin), APOA1 (Apolipoprotein A1), APOC1 (Apolipoprotein C1), GSN (Gelsolin), TF (Transferrin).

Because of the rapid action required in an early phase of a pandemic, we sampled depending on the first patients hospitalized (i.e. there was no other inclusion criteria than a hospitalisation due to a SARS-COV19 infection). Such a cohort certainly differs from a long-term planned epidemiological cohort such as the GS study. With samples collected as part of the hospital routine by different medical professionals, the level of sample variability is expected to be higher. Moreover, it is difficult to assemble a control cohort that is matched for the key confounding factors, like age. Nonetheless, our platform yielded only slightly inferior values compared to the ideal-case scenario of the GS study. In the exploratory cohort, that included 104 serum samples obtained from the 31 of the earliest COVID-19 patients, we quantified 297 protein groups among which 229 unique proteins were detected with 75% data completeness. We surmise that this somewhat lower data completeness was caused by the massive changes in levels of a number of proteins upon severe SARS-CoV-2 infection, as well as by the decrease in total serum protein content, also observed previously in patients hospitalized in ICU units (Nie et al., 2020). To account for this biological limitation, we applied very strict filtering to the dataset, namely, we only tested for differential abundance of proteins which had at least five different peptide precursors identified at least in one of the acquisitions.

We identified 37 protein groups with either increasing or decreasing levels, depending on the severity of the disease (0.05 significance, multiple testing-corrected, Theil-Sen test against the WHO severity score; see Methods for testing methodology) (Figure 4c and Table S3). Next, to validate the biomarkers, we processed the validation cohort (Table S1), and recorded 96 proteomes in triplicates for 15 healthy volunteers, and 17 COVID-19 patients (Table S1). The experiment quantified 319 protein groups among which 248 unique proteins were detected with 85% data completeness. Despite being conducted on a different matrix (citrate plasma), this independent study confirmed 27 of the protein groups with either increasing or decreasing levels (A1BG, ACTB;ACTG1, ALB, APOA1, APOC1, C1R, C1S, C8A, CD14, CFB, CFH, CFI, CRP, FGA, FGB, FGG, GSN, HP, ITIH3, ITIH4, LBP, LGALS3BP, LRG1, SAA1, SAA1;SAA2, SERPINA10, TF; 0.05 significance, multiple testing-corrected). This set of proteins thus represents potential biomarkers of disease severity. Out of the remaining 10 proteins, 9 (AGT, AZGP1, C2, C7, C8B, CLU, CPN1, PLG, VTN) did not reach statistical significance in the smaller validation group, while the IGHG2;IGHG3 protein group showed the opposite trend. We illustrate the quantitative variability of the validated biomarkers for COVID-19 severity on a heatmap (Figure 4c) and as boxplots (Figure 4d, Figure S4), as well as summarize their potential connection to COVID-19 in Table S3.

We illustrate the concentration changes of the biomarkers validated in the control cohort, upon grouping of the patients according the WHO severity criteria, ranging from Scale 3 (hospitalized, no oxygen therapy) to the most critical (Scale 7), in a heatmap (Table S1 for the grading of each patient), which graphically illustrates how level changes in these proteins reflect a progression from mild to severe COVID-19 (Figure 4c). Moreover, principal component analysis categorizes the individuals according to the severity of COVID-19 (Figure 4b).

Notably, when we first obtained the dataset, two individuals (Patients 4 and 6) initially clinically assessed as severe (Braun et al., 2020) clustered with patients that suffered from the mild form of COVID-19 (Figure S1). Our results triggered a retrospective assessment, which revealed that patient 4 turned out to be suffering from type B influenza rather than a SARS-CoV-2 infection, whereas patient 6 was classified as severe due to recent R-CHOP chemotherapy, applied just ten days before his COVID-19-related hospitalisation. We have, as a consequence, excluded Patient 4 from the COVID-19 dataset (all results shown in Figure 4), but both clinical findings indicate a high prognostic precision of the proteomic biomarker signatures.

To exclude the possibility that concentration changes in these markers are due to frequent confounders, like age, we have plotted the variability of the same proteins in the GS cohort, with age spanning from 37 to 79, on samples that have been collected before the COVID-19 outbreak. We note that for identified proteins the change between mild and severe COVID-19 substantially exceeds the variation seen in the general population (Figure 4d). Moreover, plotting the protein abundance values against the age did not reveal significant correlations across the GS population baseline (Figure S5).

### 4. Proteome signatures of inflammation and acute-phase response in severe COVID-19

We observed that most of the differentially expressed genes between COVID-19 cases of different severity are implicated in the inflammatory response and the acute phase response system. We detected consistent activation of both the classical complement pathway (C1R, C1S, C8A) as well as the alternative pathway Factor B (CFB) and the complement modulators: Factors I (CFI) and H (CFH). Other differentially expressed proteins included the common acute phase reactants, such as C-Reactive Protein (CRP, upregulated), Albumin (ALB, downregulated), or Serum Amyloid proteins SAA1 and SAA2 (upregulated).

We observed upregulation of a number of proteins implicated in interleukin IL-6 signalling (Figures 4 and 5, Table S3). In addition to SAA1 and SAA2, these include Inter-*α*-Trypsin Inhibitor Heavy Chain 4 (ITIH4), which plays an important role in extracellular matrix organization and is implicated in inflammation (Bost et al., 1998; Yang et al., 2012), haptoglobin (HP), an acute-phase response protein (Jain et al., 2011), Leucine-Rich Alpha-2-Glycoprotein (LRG1), a promoter of cell proliferation and angiogenesis, implicated in local inflammation and fibrosis (Honda et al., 2017), Monocyte Differentiation Antigen CD14, primarily involved in bacterial LPS recognition (Kielian and Blecha, 1995) and the Liposaccharide Binding Protein (LBP), as well as Galectin 3 Binding Protein (LGALS3BP), a pro-inflammatory factor, which is known to induce IL-6 expression (Silverman et al., 2012).

We also observe upregulation of fibrinogen, a coagulation factor, and Protein Z-Dependent Protease Inhibitor, SERPINA10, an inhibitor of the F10a coagulation factor, further highlighting the importance of coagulation in SARS-CoV-2 infection, established by previous observations of elevated coagulation in severe COVID-19 cases (Zhou et al., 2020a).

A recent study (Shen et al., 2020) used a more conventional and time consuming (120x pre-fractionation consolidated in 40 fractions with TMT-16 plex) proteomics method to characterize plasma samples from 99 study participants, including 46 samples from patients with COVID-19 diagnosed in China. Despite the different cohorts and different technologies used, the proteomes implicate similar biological mechanisms in the differentiation of mild, severe or critical disease progression. Many of the proteins that differentiate the groups in both studies belong to the complement system, acute phase and inflammatory response. For example, both studies agree with independently conducted clinical investigations on a number of differentially expressed proteins, in particular, albumin, the complement factors, Serum Amyloid proteins, ITIH3 and ITIH4 (Nie et al., 2020; Shen et al., 2020). Despite these similarities, we note important differences. For instance, we cannot confirm the downregulation of Pro-Platelet Basic Protein (PPBP) and Platelet Factor 4 (PF4) in severe COVID-19 as highlighted by Shen et al. We can offer several potential explanations. We note that different SARS-CoV-2 clades might exhibit different degrees of pathogenicity (Yao et al., 2020) and thus elicit different physiological responses, especially in different populations. However, when examining the response of PF4 and PPBP at the peptide level, we discovered that while several peptides maintain a relatively stable level, other PF4- and PPBP-specific peptides increase or decrease in concentration (Figure S2). This situation might indicate that instead of being differentially expressed, PF4 and PPBP might be differentially post-translationally modified.

### 5. Plasma proteomes provide insights into the virulence mechanisms and potential therapeutic targets for COVID-19

Extensive worldwide efforts have been directed recently into finding drug targets for COVID-19 treatment. As most of the damage associated with severe SARS-CoV-2 infection appears to be indirect and caused by excessive inflammation in the lungs, it is of crucial importance to seek opportunities not only to target the pathway of entry and the replication mechanism of the virus, but to also identify and examine the possibilities for targeting host factors responsible for harmful inflammatory responses, to both alleviate the severity of the infection and to reduce the chance of long-lasting complications (Huang et al., 2020). Some preliminary results in that direction appear promising. For example, proinflammatory signalling via interleukin IL-6 has been determined to be a marker of severe COVID-19 (Chen et al., 2020; Ruan et al., 2020) and preliminary results on the inhibition of IL-6 receptor (IL-6R) with tocilizumab seem to indicate clinical improvement (Coomes and Haghbayan, 2020). Here we highlight several novel biomarkers of severe COVID-19 that are linked to IL-6-mediated proinflammatory cytokine signalling: i) the CD14-LBP LPS-recognition system, ii) upregulation of Leucine-rich-alpha-2-glycoprotein 1 (LRG1), an angiogenesis and anti-apoptotic factor associated with inflammation, iii), upregulation of the Galectin 3 Binding Protein (LGALS3BP), an inducer of IL-6. Below we discuss the potential significance of these findings.

Coronaviruses are known to actively disrupt the host immune response (Enjuanes et al., 2016). For example, their papain-like proteases (PLPs) act as interferon antagonists (Niemeyer et al., 2018), causing delayed type-I interferon response, macrophage-mediated inflammation and lung damage (Channappanavar et al., 2016). Here we observed upregulation of both Monocyte Differentiation Antigen CD14 (~2.7x) and Lipopolysaccharide Binding Protein (LBP; ~3.9x) in severe COVID-19. As the response to bacterial LPS is one of the primary functions of both CD14 and LBP, which can act in complex to sensitise toll-like receptor-mediated LPS recognition (Ranoa et al., 2013), this observation reflects the dysregulation of the innate immune response by SARS-CoV-2, leading to the activation of the anti-bacterial defense and sensitisation to LPS, thus contributing to excessive inflammation, with effects likely more pronounced in case of a concomitant secondary bacterial infection. Interestingly, CD14 and LBP upregulation has been observed in viral pneumonia before (Van Gucht et al., 2005), while CD14 is one of the primary mediators of lung inflammation (Anas et al., 2010). Of note, the GPI-anchored form of CD14 is primarily displayed by monocytes and macrophages (Marcos et al., 2010), while the proportion of CD14^+^CD16^+^ inflammatory monocytes in the peripheral blood increases along with COVID-19 severity (Zhou et al., 2020b). At the same time, IL-6 induces soluble CD14 production in the liver (Bas et al., 2004), as well as, along with other cytokines, release of CD14 from monocytes upon their activation (Shive et al., 2015). Given that CD14 is a potent activator of proinflammatory cytokine signalling (Zanoni and Granucci, 2013), it might present a potential therapeutic target for COVID-19.

LRG1 is another pro-inflammatory factor induced by IL-6 (Shirai et al., 2009), which is known to promote angiogenesis and cell proliferation, while inhibiting apoptosis (Meng et al., 2016; Naka and Fujimoto, 2018; Wang et al., 2013). Some recent findings indicate its role in promoting skin fibrosis and lung fibrosis in Transforming Growth Factor beta (TGF-β)-mediated fashion (Gao et al., 2019; Honda et al., 2017). Given the previously reported association of the MERS infection with lung fibrosis (Zhao et al., 2008) and emerging reports (British Thoracic Society, 2020) of lung fibrosis in a substantial proportion of COVID-19 survivors, we hypothesize that the ~2.1x elevation of serum LRG1 levels we observe in critical COVID-19 cases in comparison to the mild cases might indicate the increased risk of fibrosis, highlighting LRG1 as yet another potential therapeutic candidate for COVID-19 treatment. Furthermore, we detected about ~3.4x upregulation of LGALS3BP, which is known to induce the expression of IL-6 by stromal cells in Galectin-3-dependent manner (Silverman et al., 2012). Of note, Galectin-3 has long been considered an attractive drug target in combating various forms of TGF-β-mediated fibrosis and pathological inflammatory conditions (Brinchmann et al., 2018; Mackinnon et al., 2012; Shen et al., 2018; Yu Lili et al., 2013). Inhibition of Galectin-3-mediated signalling pathways hence represents another potential therapeutic target against COVID-19.

### 6. Tissue injury and dysregulation of modulators of inflammation

We report substantially decreased (~2.6x) levels of gelsolin (GSN) (Figure 4b,c). Plasma gelsolin is a part of the extracellular actin scavenger system (EASS), which removes toxic F-actin filaments that have been released from necrotic cells to the bloodstream (Piktel et al., 2018). Low levels of plasma gelsolin are associated with inflammation: it is believed that gelsolin is recruited to the sites of tissue injury to handle the released actin, depleting its plasma levels. Interestingly, we do observe the increase in serum actin concentration (Beta and Gamma-1 actin, ~2.2x), indicative of tissue injury (DiNubile, 2008), which could explain the gelsolin depletion from the blood. Importantly, plasma gelsolin is a powerful modulator of inflammation, which carries a protective function (DiNubile, 2008; Li et al., 2012). Low plasma gelsolin is a marker of poor prognosis in various pathological conditions, including diabetes (Khatri et al., 2014), cancers (Asare-Werehene et al., 2019; Stock et al., 2015) and sepsis (Lee et al., 2007), leading to suggestions and animal tests for its therapeutic use. Going forward, it will be important to assess GSN levels in at-risk populations for severe COVID-19, e.g. patients with diabetes. Of note, treatment with gelsolin has been observed to decrease IL-6 levels in mice (Cheng et al., 2017) and has been suggested to promote epithelial repair (Wittmann et al., 2018). The development of therapies to stabilise the gelsolin levels could hence be of direct therapeutic value for treating COVID-19.

Notably, we observed a decrease in the expression levels of Apolipoprotein A1 (APOA1; ~3x). APOA1 is a major component of the High-Density Lipoprotein (HDL) complex, which is a modulator of innate immune response and inflammation (Fotakis et al., 2019; Gordon et al., 2011; Macpherson et al., 2019; White et al., 2017). We also observe decreased levels of Apolipoprotein C1 (~3.2x), a component of several lipoprotein complexes (Fuior and Gafencu, 2019). Based on the GS study, we note that the serum levels of APOA1 are correlated with those of HDL cholesterol (Figure S3). Although decreased APOA1 levels have been observed in systemic inflammatory response (Kumaraswamy et al., 2012; Sirniö et al., 2017), including in COVID-19 (Nie et al., 2020), a potential explanation of the downregulation we observe here would also be provided, if naturally lower APOA1, and hence a different metabolic condition of the individual, were associated with a higher risk of severe SARS-CoV-2 disease progression.

In conclusion, SARS-CoV-2, SARS and MERS constitute a class of emerging coronaviruses of high public health concern. It is likely that other viruses will emerge in the future for which at time of outbreak insufficient biochemical knowledge will be available to identify biomarkers and to define point of care clinical classifiers. Serum proteomics can present valuable and unbiased information about disease progression and therapeutic candidates, without prior knowledge about the etiologies and biomolecules involved. We present a new workflow for rapid and large-scale clinical proteomics that is completely redesigned in comparison to previous platforms. The sample preparation workflow scales to high sample numbers, enables high quantification precision, and reduces batch effects for large-scale and longitudinal studies, while the data acquisition and processing workflow is able to exploit the advantages of high-flow chromatography in short-gradient proteomics, which improves throughput, data quality, and greatly simplifies implementation in regulated laboratories. We demonstrate a quantification precision and acquisition robustness that, to our knowledge, has not previously been shown in large-scale proteomic experiments. We then applied the technology to a cohort of early hospitalized cases of the SARS-CoV-2 pandemic. We identify biomarkers that distinguish mild and severe forms of COVID-19, highlighting the role of complement factors, the coagulation system, several inflammation modulators as well as pro-inflammatory signalling both upstream and downstream of interleukin IL-6. The proteomic signatures and biomarkers identified pave the way for the development of routine assays to support clinical decision making, as well as provide new hypotheses about potential COVID-19 therapeutic targets.

## Methodology

### Materials

Water (LC-MS Grade, Optima; 10509404), Acetonitrile (LC-MS Grade, Optima; 10001334) and Methanol (LC-MS Grade, Optima, A456–212) were purchased from Fisher Chemicals. DL-Dithiothreitol (Biollltra, 43815), lodoacetamide (Biollltra, 11149) and Ammonium Bicarbonate (Eluent additive for LC-MS, 40867) were purchased from Sigma Aldrich. Urea (puriss. P.a., reag. Ph. Eur., 33247H) and Acetic Acid (Eluent additive for LC-MS, 49199) were purchased from Honeywell Research Chemicals. Trypsin (Sequence grade, V511X) and K562 cell lysate (MS Compatible Human Protein Extract, Digest, V6951) were purchased from Promega. iRT peptides (Ki-30002-b) were purchased from Biognosys. Control samples for SARS-CoV-2 study were prepared from Human Serum (Sigma Aldrich, S7023–50MB) and Human Plasma (EDTA, Pooled Donor, Genetex GTX73265). Control samples for GS study were prepared from Human Serum (tebu-bio, 088HSER) and Human Plasma (088SER-K2EDTA). MS synthetic peptide calibration kit (5045759) was purchased from SCIEX.

### Clinical samples of COVID-19 patients

Sampling was performed as part of the Pa-COVID-19 study, a prospective observational cohort study assessing pathophysiology and clinical characteristics of patients with COVID-19 at Charité Universitätsmedizin Berlin (Braun et al., 2020). All patients with SARS-CoV-2 infection proven by positive PCR from respiratory specimens and willing to provide written informed consent are eligible for inclusion. Exclusion criteria are refusal to participate in the clinical study by patient or legal representative or clinical conditions that do not allow for blood sampling. The study assesses epidemiological and demographic parameters, medical history, clinical course, morbidity and quality of life during hospital stay of COVID-19 patients. Moreover, serial high-quality bio-sampling consisting of various sample types with deep molecular, immunological and virological phenotyping is performed. Treatment and medical interventions follow standard of care as recommended by current international and German guidelines for COVID-19. Severity of illness in the present study follows the WHO ordinal outcome scale (Tables S1, S2). The Pa-COVID-19 study is carried out according to the Declaration of Helsinki and the principles of Good Clinical Practice (ICH 1996) where applicable and was approved by the ethics committee of Charité- Universitätsmedizin Berlin (EA2/066/20).

### Generation Scotland study

199 serum samples from random individuals that participated in the Generation Scotland (GS) epidemiological study (Smith et al., 2013) were used. GS is a family-based cohort of approximately 24,000 individuals in 7,000 family groups from across Scotland, aged between 18 and 98 (Smith et al., 2013). All components of Generation Scotland received ethical approval from the NHS Tayside Committee on Medical Research Ethics (REC Reference Number: 05/S1401/89). All participants provided broad and enduring written informed consent for biomedical research. Generation Scotland has also been granted Research Tissue Bank status by the East of Scotland Research Ethics Service (REC Reference Number: 15/0040/ES), providing generic ethical approval for a wide range of uses within medical research. This study was performed in accordance with the Helsinki declaration.

### Plasma and serum sample preparation

The protocol was designed for preparing four 96-well plates in parallel. All liquid transfer and mixing except the addition of serum/plasma to the denaturing buffer was carried out by the liquid handling robot, either a Beckman Coulter Biomek NPx (Crick laboratory) or Biomek i7 liquid handling robot (Charité Universitätsmedizin Berlin). There are slight differences between the protocols due to the two different laboratories. To our knowledge, these have no detectable influence on the results. Where applicable these differences are indicated as “Biomek NPx” or “Biomek i7 protocol”, respectively.

Before starting the sample preparation, 96-well plates were prefilled with Trypsin (12.5μl, 0.1μg/|μl solution; four plates/batch), denaturation/reduction buffer (55μl 8M Urea, 100mM ammonium bicarbonate (ABC) and 4.5mM dithiothreitol (DTT) (Biomek NPx protocol) or 50mM DTT (Biomek i7 protocol); four plates/batch) and iodoacetamide (IAA) (100mM, > 20 μl, one plate/batch) and stored at −80°C until the day of the experiment.

5μl of thawed serum/plasma samples were transferred to the pre-made denaturation/reduction stock solution plates. Subsequently the plates were centrifuged for 15s at pulse setting (Eppendorf Centrifuge 5810R), mixed and incubated at 30°C for 60 minutes. The mixing in this step was done either 30s at 1000rpm on a Thermomixer (Eppendorf Thermomixer C) (Biomek NPx protocol) or by resuspension (Biomek i7 protocol). 5μl IAA was then transferred from the respective stock solution plate to the sample plate and incubated in the dark at 23°C for 30 minutes before dilution with 100mM ABC buffer (340μl). 220μl of this solution was transferred to the pre-made trypsin stock solution plate and incubated at 37°C for 17 h. The digestion was quenched by addition of formic acid (10% v/v, 25μl). The digestion mixture was cleaned-up using C18 96-well plates (BioPureSPE Macro 96-Well, 100mg PROTO C18, The Nest Group). For the solid phase extraction, 1 minute of centrifugation at the described speeds (Eppendorf Centrifuge 5810R) was used to push the liquids through the stationary phase and the liquid handler was used to pipette the liquids onto the material in order to make four 96-well plates/batch feasible. The plates were conditioned with methanol (200μl, centrifuged at 50g), washed twice with 50% ACN (200μl, centrifuged at 150g and flow through discarded), equilibrated twice with with 0.1% FA (200μl, centrifuged at 150g and flow through discarded). Then 200μl digested and quenched samples were loaded (centrifuged at 150g), washed twice with 0.1% FA (200μl, centrifuged at 150g). After the last washing step, the plates were centrifuged another time at 200g before the peptides were eluted in 3 steps with 110μl 50% ACN (200g) into a new collection plate (1.1ml, Square well, V-bottom). Collected material was completely dried on a vacuum concentrator (Eppendorf Concentrator Plus (Biomek NPx protocol) or Fisher Scientific, SPD300P1 (Biomek i7 protocol) and redissolved in 50μl 1% ACN, 0.1% formic acid (Biomek NPx protocol) or 50μl 0.1% formic acid (Biomek i7 protocol), then stored at −80°C until data acquisition. The samples for the SARS-CoV-2 studies were directly analysed. QC samples for repeat injections were prepared by pooling commercial serum samples and were spiked with iRT peptides (Biognosys).

### Liquid chromatography-mass spectrometry setup

Liquid chromatography was established on on two complementary and exchangeable ultra-high-pressure highflow LC-MS systems, an Agilent 1290 Infinity II (Crick laboratory) and Waters H-Class (Charité Universitätsmedizin Berlin) system, both coupled to a TripleTOF 6600 mass spectrometer (SCIEX) equipped with IonDrive Ion Source (Sciex). In both cases, the peptides were separated in reversed phase mode using a C18 ZORBAX Rapid Resolution High Definition (RRHD) column 2.1mm × 50mm, 1.8 μm particles. A linear gradient was applied which ramps from 3% B to 36% B in 5 minutes (Buffer A: 0.1% FA; Buffer B: ACN/0.1% FA) with a flow rate of 800μl/min. For washing the column, the organic solvent was increased to 80% B in 0.5 minutes and was kept for 0.2 minutes at this composition before going back to 1% B in 0.3 min. The equilibration times were 2.8 minutes (Water H Class protocol) or 4.2 minutes (Agilent Infinity II protocol). Data was acquired in high sensitivity mode and the amount of total proteins injected was 5 μg (GS study) and 10 μg (SARS-CoV-2). The DIA/SWATH method consisted of an MS1 scan from m/z 100 to m/z 1500 (20ms accumulation time) and 25 MS2 scans (25ms accumulation time) with variable precursor isolation width covering the mass range from m/z 450 to m/z 850. An IonDrive Turbo V Source (Sciex) was used with ion source gas 1 (nebulizer gas), ion source gas 2 (heater gas) and curtain gas set to 50, 40 and 25 respectively. The source temperature was set to 450 and the ion spray voltage to 5500V.

### Mass spectrometry data processing, batch correction and and quality control

The raw data were processed using DIA-NN 1.7.10 in high-precision mode with RT-dependent median-based cross-run normalisation enabled. MS2 and MS1 mass accuracies were set to 20 and 12 ppm, respectively, and scan window size set to 6. Although DIA-NN can optimise such parameters automatically, we fixed them to these values to ensure comparability. For each of the experiments, we used a project independent, public spectral library (Bruderer et al., 2019). Spectra and retention times were first automatically refined based on the dataset in question at 0.01 q-value (using the “Generate spectral library” option in DIA-NN) and the refined library was then used to reanalyse the data. The resulting report was stringently filtered at 0.01 precursor-level q-value, 0.005 precursor-level library q-value and 0.05 protein group-level q-value. Intra-batch correction was performed on log-transformed peptide precursor quantities using linear regression on the total injection number based on replicate injections, or running median smoothing, whatever performed best. Linear regression was applied only for at least 10 data points and if the p-value for non-zero slope was below 0.01. Running median smoothing was performed in two steps. First, a 5-point smoothing for the control samples, to remove outliers. Second, interpolation of the resulting values to all injections followed by 41-point smoothing. In “GS”, no plate-specific bias was detected and thus no plate-specific correction was applied. Protein quantification was performed using the MaxLFQ algorithm (Cox et al., 2014) as implemented in the diann R package (https://github.com/vdemichev/diann-rpackage. version 1.0, commit “eb4607a”). Data completeness was defined as the proportion of non-missing values in the proteins x samples quantities matrix. The coefficient of variation (CV) was calculated for each protein as its empirical standard deviation divided by its empirical mean. PCA analysis was always performed only on ubiquitously identified proteins: imputation was not used.

### Differential expression analysis

In the exploratory cohort, differential expression was tested only for proteins quantified using at least five peptide precursors in one of the acquisitions. Further, protein groups were quantified using only precursors detected in at least 10% of patient samples (both cohorts). As we were specifically interested in proteins which could serve as biomarkers of COVID-19 severity, the test was performed using the Kendall’s Tau test for the Theil-Sen trend estimator (as implemented in the EnvStats R package (Millard, 2014)) against the disease severity as classified according to the WHO ordinal scale (Table S1). The input for the test was obtained by calculating, for each patient, median protein levels across the timepoints or replicates measured. Significance threshold was set to 0.05 (multiple testing-corrected). The choice of a nonparametric test (Theil-Sen) was dictated by the fact that such widespread methods as ANOVA or linear regression are only valid under the assumption of Gaussian errors with the same variance across all conditions. In the case of this dataset, however, we observed very significant differences in the variance, e.g. many proteins seem a lot more variable between patients with severe and critical COVID-19 than mild COVID-19. For such a situation a nonparametric test is ideal: although it would typically have less power (less proteins detected as differentially expressed), the p-values produced are reliable.

### Chromatographic peaks full width at half maximum (FWHM) estimations

Median peak FWHM was estimated using Spectronaut 13 (version 13.12.200217.43644; Biognosys). Only precursors ubiquitously identified in all runs (3 minutes, 5 minutes, 10 minutes and 20-minute high-flow as well as with the 20-minute micro-flow run) and with a q-value of < 0.001 were considered (804 precursors total).

### Total ion and extracted ion chromatograms

Total ion chromatograms and extracted ion chromatograms were generated with the PeakView software (Version 2.2, SCIEX), exported and plotted in R (R core team, www.R-project.org).

### Data availability

The raw data of the acquired commercial plasma and serum control samples within the GS study was submitted to the ProteomeXchange Consortium via PRIDE (Perez-Riverol et al., 2019) partner repository with the dataset submission #413758. According to the terms of consent for GenerationScotland participants, access to individual-level data (omics and phenotypes) must be reviewed by the GS Access Committee. Applications should be made to.

## Acknowledgements

We thank Stephan Kamrad and Simran Aulakh for proofreading the manuscript, Jan-David Manntz (Beckman, Germany) for help with the Biomek i7, Robert Lane, Jean-Baptiste Vincedent and Nick Morrice (SCIEX) for help with the TripleTOF 6600. This work was supported by the Ministry of Education and Research (BMBF), as part of the National Research Node ‘Mass spectrometry in Systems Medicine (MSCoresys), under grant agreement 031L0220A. The study was further supported by the Francis Crick Institute, which receives its core funding from Cancer Research UK (FC001134), the UK Medical Research Council (FC001134), and the Wellcome Trust (FC001134), and received specific funding from the BBSRC (BB/ N015215/1 and BB/N015282/1) and the Wellcome Trust (200829/Z/16/Z) as well as a Crick Idea to Innovation (i2i) initiative (grant number 10658) (to MR). The Generation Scotland study received core support from the Chief Scientist Office of the Scottish Government Health Directorates (CZD/16/6) and the Scottish Funding Council (HR03006), and is now supported by the Wellcome Trust (216767/Z/19/Z). Archie Campbell is funded by HDR UK and the Wellcome Trust (216767/Z/19/Z). Caroline Hayward is supported by an MRC University Unit Programme Grant (MC_UU_00007/10) (QTL in Health and Disease). Riccardo Marioni is supported by an Alzheimer’s Research UK project grant (ARUK-PG2017B-10). Leif Erik Sander is supported by the German Research Foundation (DFG, SFB-TR84 114933180) and by the Berlin Institute of Health (BIH), which receives funding from the Ministry of Education and Research (BMBF).

## Supplementary information

**Table S1.** Patient information table for the exploratory and validation COVID-19 cohorts.

| Exploratory Cohort |  |  |  |  |  |  |
| --- | --- | --- | --- | --- | --- | --- |
| Patient | WHO score | WHO category | Clinical assessment (Charité) | Age | Sex | Outcome |
| 1 | 3 | mild | mild | 21 | m | discharged |
| 2 | 3 | mild | mild | 31 | f | discharged |
| 3 | 3 | mild | mild | 45 | m | discharged |
| 10 | 3 | mild | mild | 50 | f | discharged |
| 18 | 3 | mild | mild | 60 | f | discharged |
| 19 | 3 | mild | mild | 52 | m | discharged |
| 23 | 3 | mild | mild | 44 | f | discharged |
| 27 | 3 | mild | mild | 40 | f | discharged |
| 28 | 3 | mild | mild | 64 | m | discharged |
| 34 | 3 | mild | mild | 47 | m | discharged |
| 35 | 3 | mild | mild | 37 | f | discharged |
| 37 | 3 | mild | mild | 78 | m | discharged |
| 4* |  |  | severe* | 37 | m |  |
| 6 | 3 | mild | severe | 24 | m | discharged |
| 5 | 4 | mild | severe | 71 | m | discharged |
| 7 | 4 | mild | severe | 32 | m | discharged |
| 24 | 4 | mild | severe | 64 | m | discharged |
| 29 | 4 | mild | severe | 53 | m | discharged |
| 11 | 5 | severe | severe | 61 | m | discharged |
| 16 | 5 | severe | severe | 78 | m | still hospitalized |
| 22 | 5 | severe | severe | 62 | m | discharged |
| 26 | 5 | severe | severe | 56 | f | discharged |
| 12 | 6 | severe | critical | 75 | m | discharged |
| 20 | 6 | severe | critical | 54 | f | discharged |
| 21 | 6 | severe | critical | 52 | m | discharged |
| 8 | 7 | severe | critical | 63 | m | still hospitalized |
| 9 | 7 | severe | critical | 80 | f | still hospitalized |
| 13 | 7 | severe | critical | 71 | m | death |
| 14 | 7 | severe | critical | 81 | m | death |

| 15 | 7 | severe | critical | 54 | m | death |
| --- | --- | --- | --- | --- | --- | --- |
| 25 | 7 | severe | critical | 62 | f | still hospitalized |
| 32 | 7 | severe | critical | 50 | f | death |
| **Validation Cohort** |  |  |  |  |  |  |
| Individual/<br>Patient | WHO score | WHO<br>category | Clinical<br>category | Age | Sex | Outcome |
| HD-1 | 0 | uninfected | healthy | 27 | f |  |
| HD-2 | 0 | uninfected | healthy | 23 | m |  |
| HD-3 | 0 | uninfected | healthy | 34 | f |  |
| HD-4 | 0 | uninfected | healthy | 31 | f |  |
| HD-5 | 0 | uninfected | healthy | 30 | m |  |
| HD-6 | 0 | uninfected | healthy | 41 | f |  |
| HD-7 | 0 | uninfected | healthy | 24 | f |  |
| HD-8 | 0 | uninfected | healthy | 27 | m |  |
| HD-9 | 0 | uninfected | healthy | 30 | f |  |
| HD-10 | 0 | uninfected | healthy | 34 | f |  |
| HD-11 | 0 | uninfected | healthy | 42 | m |  |
| HD-12 | 0 | uninfected | healthy | 30 | f |  |
| HD-13 | 0 | uninfected | healthy | 33 | f |  |
| HD-14 | 0 | uninfected | healthy | 29 | f |  |
| HD-15 | 0 | uninfected | healthy | 36 | f |  |
| 49 | 3 | mild | mild | 72 | f | discharged |
| 56 | 3 | mild | mild | 58 | f | discharged |
| 65 | 3 | mild | mild | 22 | m | discharged |
| 66 | 3 | mild | mild | 75 | f | discharged |
| 75 | 3 | mild | mild | 84 | m | discharged |
| 38 | 4 | mild | severe | 70 | m | discharged |
| 48 | 4 | mild | severe | 48 | m | discharged |
| 50 | 4 | mild | severe | 78 | m | discharged |
| 63 | 5 | severe | severe | 61 | m | discharged |
| 57 | 6 | severe | critical | 55 | f | discharged |
| 60 | 6 | severe | critical | 65 | m | discharged |
| 62 | 6 | severe | critical | 72 | m | discharged |
| 33 | 7 | severe | critical | 35 | f | Still hospitalized |
| 58 | 7 | severe | critical | 26 | m | Still hospitalized |
| 59 | 7 | severe | critical | 86 | m | death |
| 61 | 7 | severe | critical | 54 | m | Still hospitalized |
| 64 | 7 | severe | critical | 57 | m | Still hospitalized |

\*A secondary assessment triggered by blind proteome clustering (Figure S1) revealed a Influenza Type B rather than a COVID-Infection.

**Table S2.**
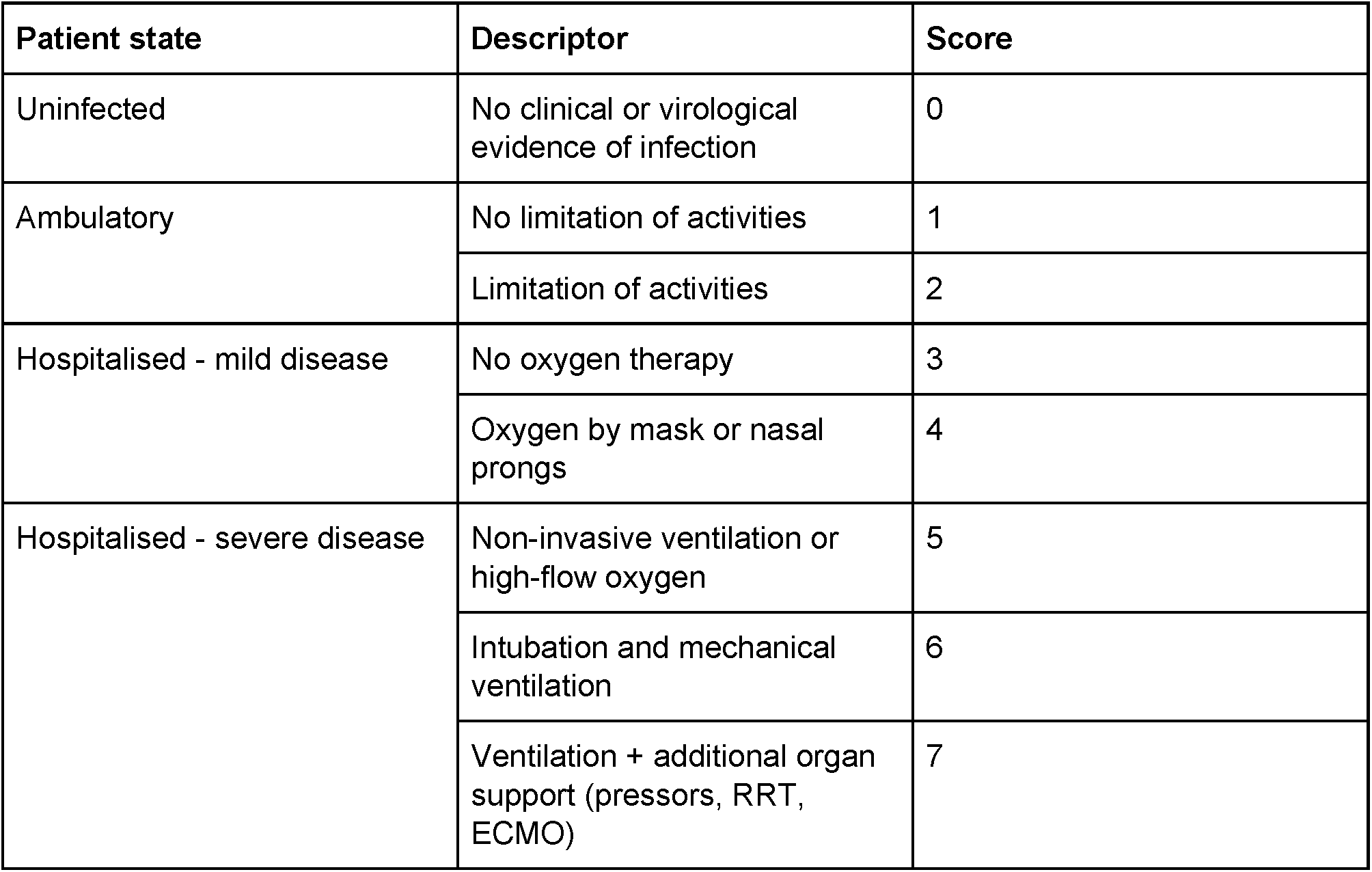
WHO scoring for COVID-19 cases, used in the study (World Health Organisation 2020).

**Table S3.**
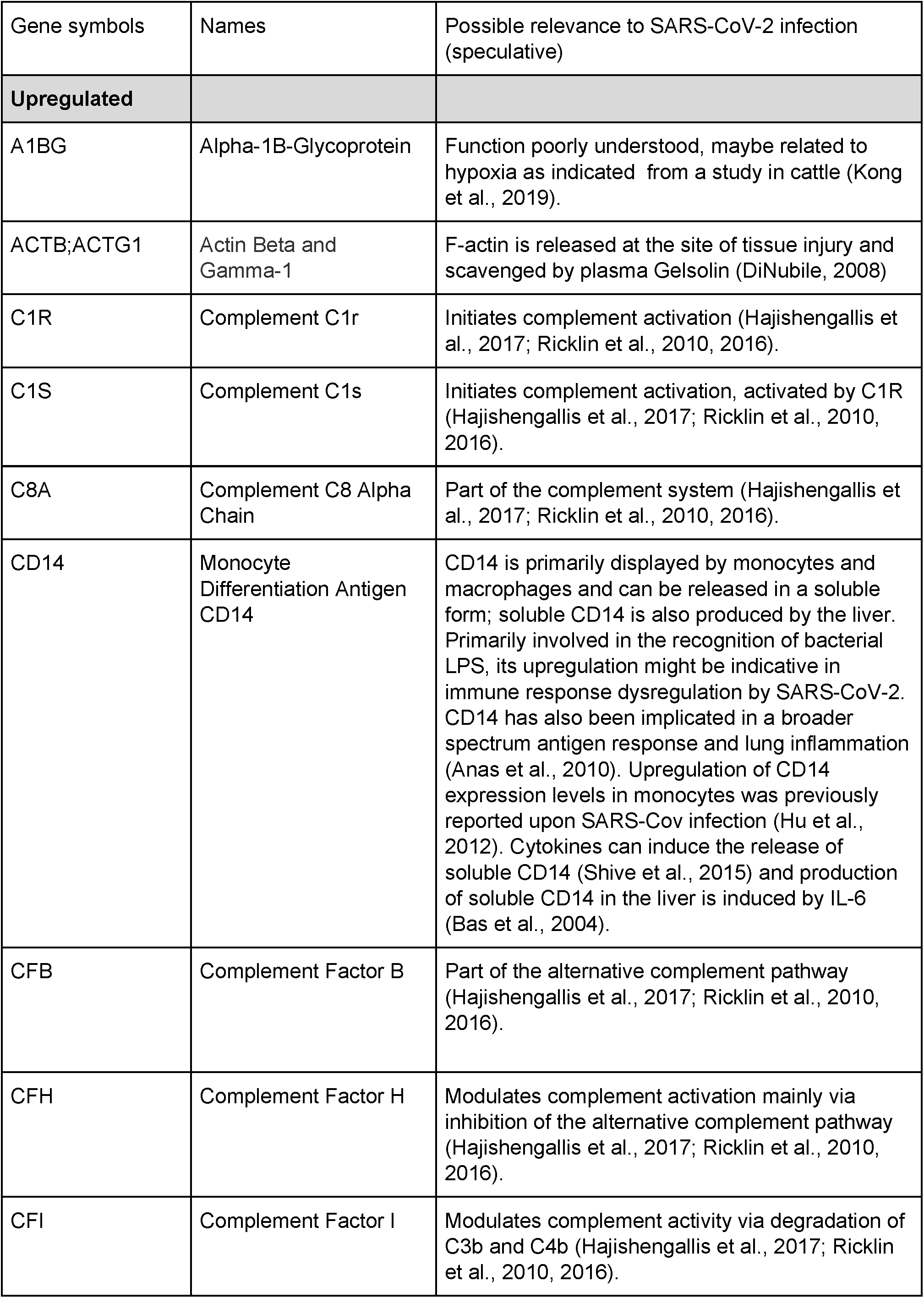
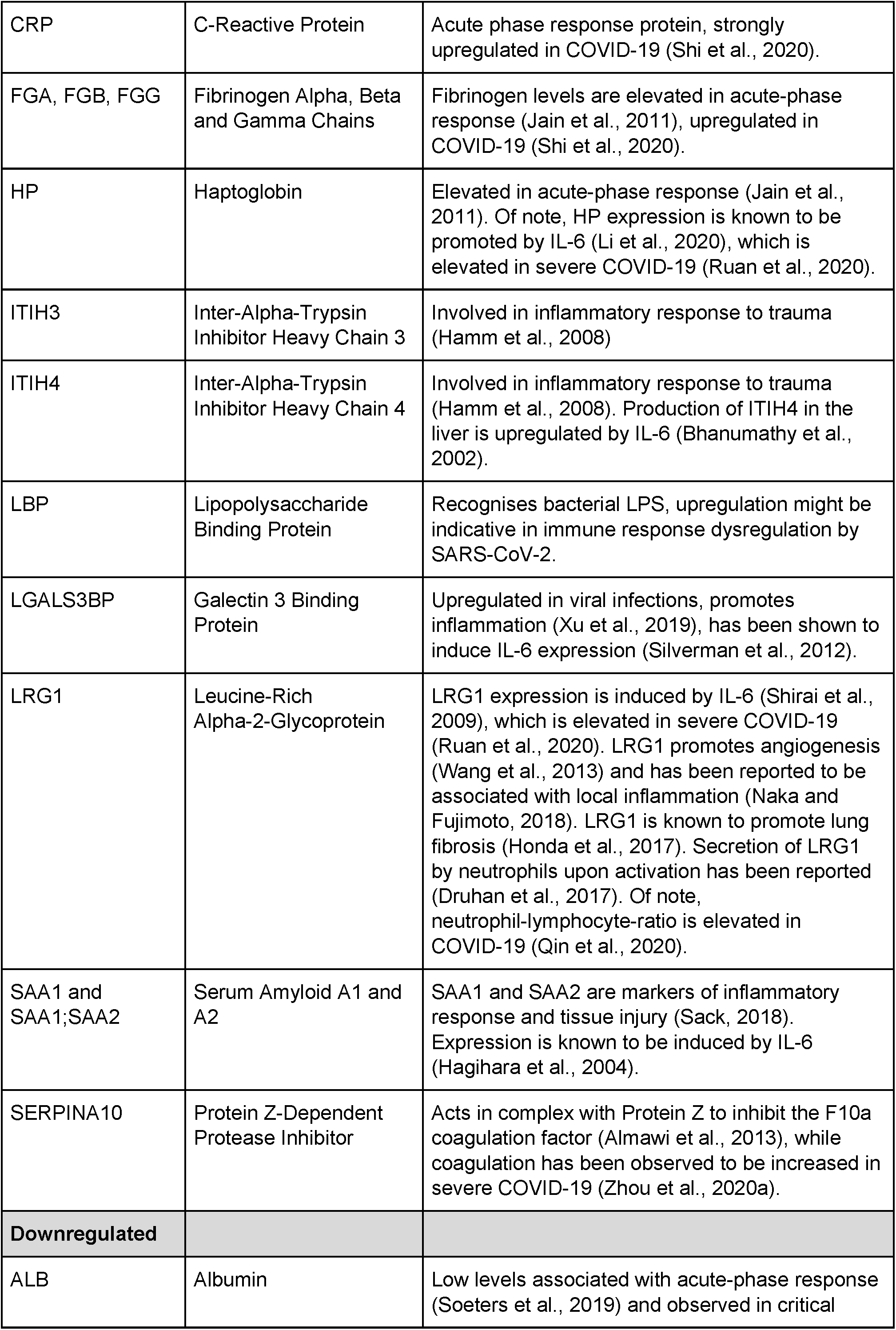
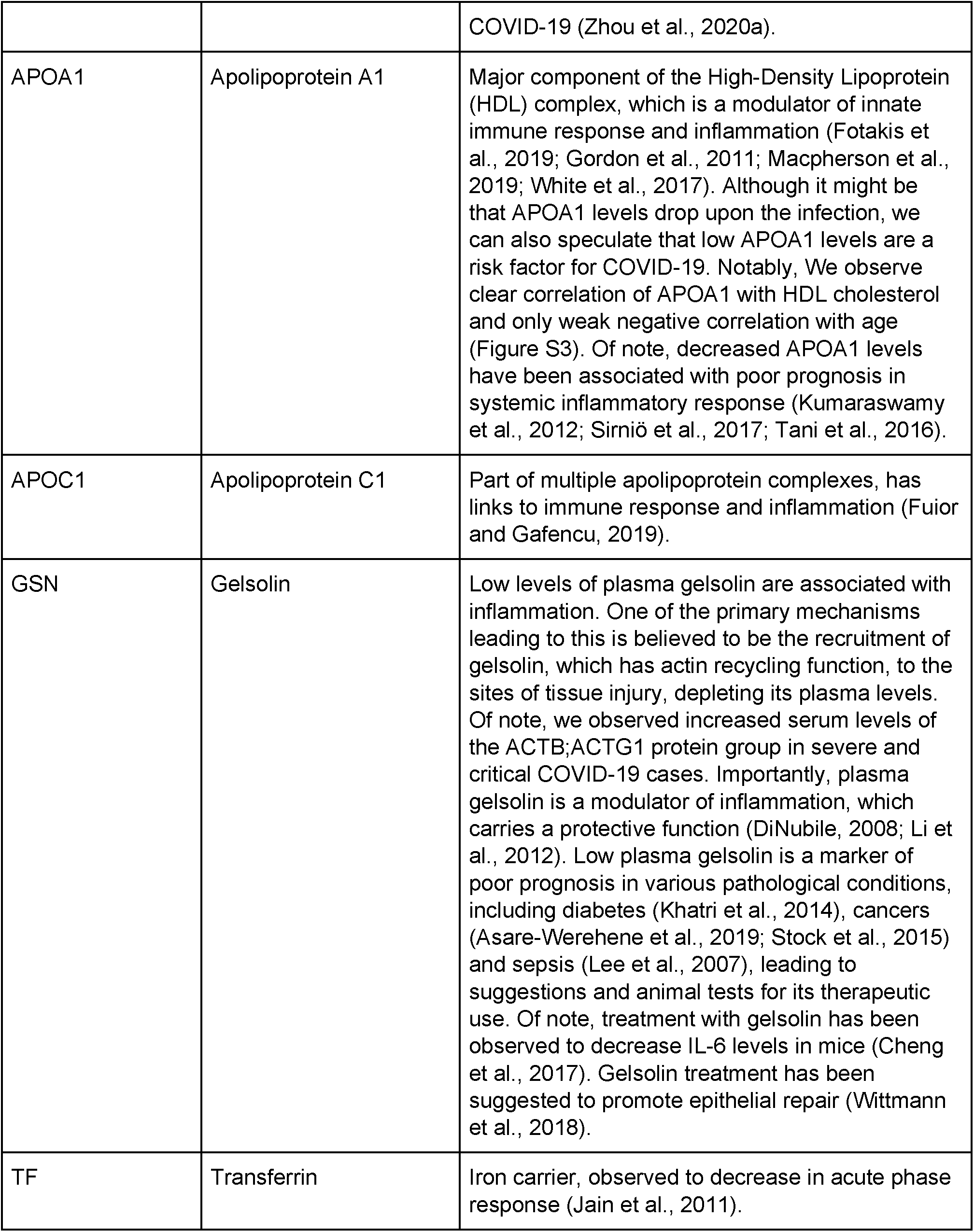
Proteins differentially expressed depending on COVID-19 severity.

**Figure S1:**
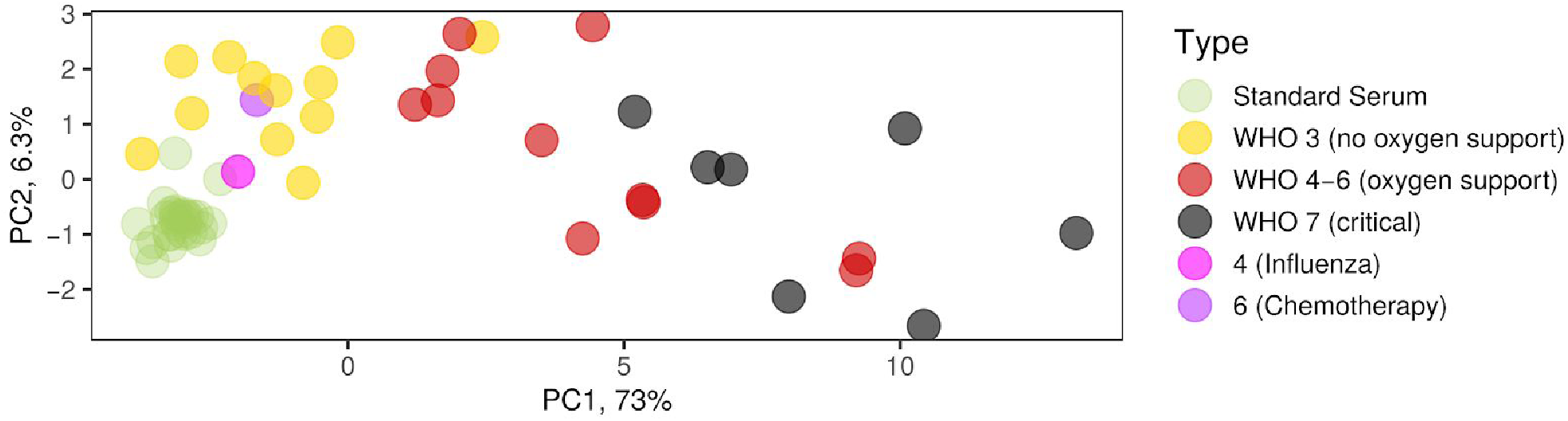
Proteome clustering led to reclassification of two study participants. Principal component analysis based on genes differentially expressed depending on COVID-19 severity grading according to WHO. Patients 4 and 6 were initially clinically assessed as “severe” (Braun et al, 2020) but clustered with patients that suffered from a mild form of COVID-19. A retrospective assessment revealed that patient 6 had received R-CHOP chemotherapy 10 days prior to study inclusion and patient 4 was in fact suffering from type B influenza rather than SARS-CoV-2 infection. Patient 6 was categorized as mild by using the WHO ordinal outcome scale of clinical improvement (World Health Organisation, 2020) and Patient 4 was removed from the analysis. Please note that the PCA transformation is slightly different from that in Figure 4b, as the addition of two extra samples for patient 4 led to slightly different protein quantities being calculated for all the samples by the MaxLFQ algorithm (Cox et al., 2014).

**Figure S2.**
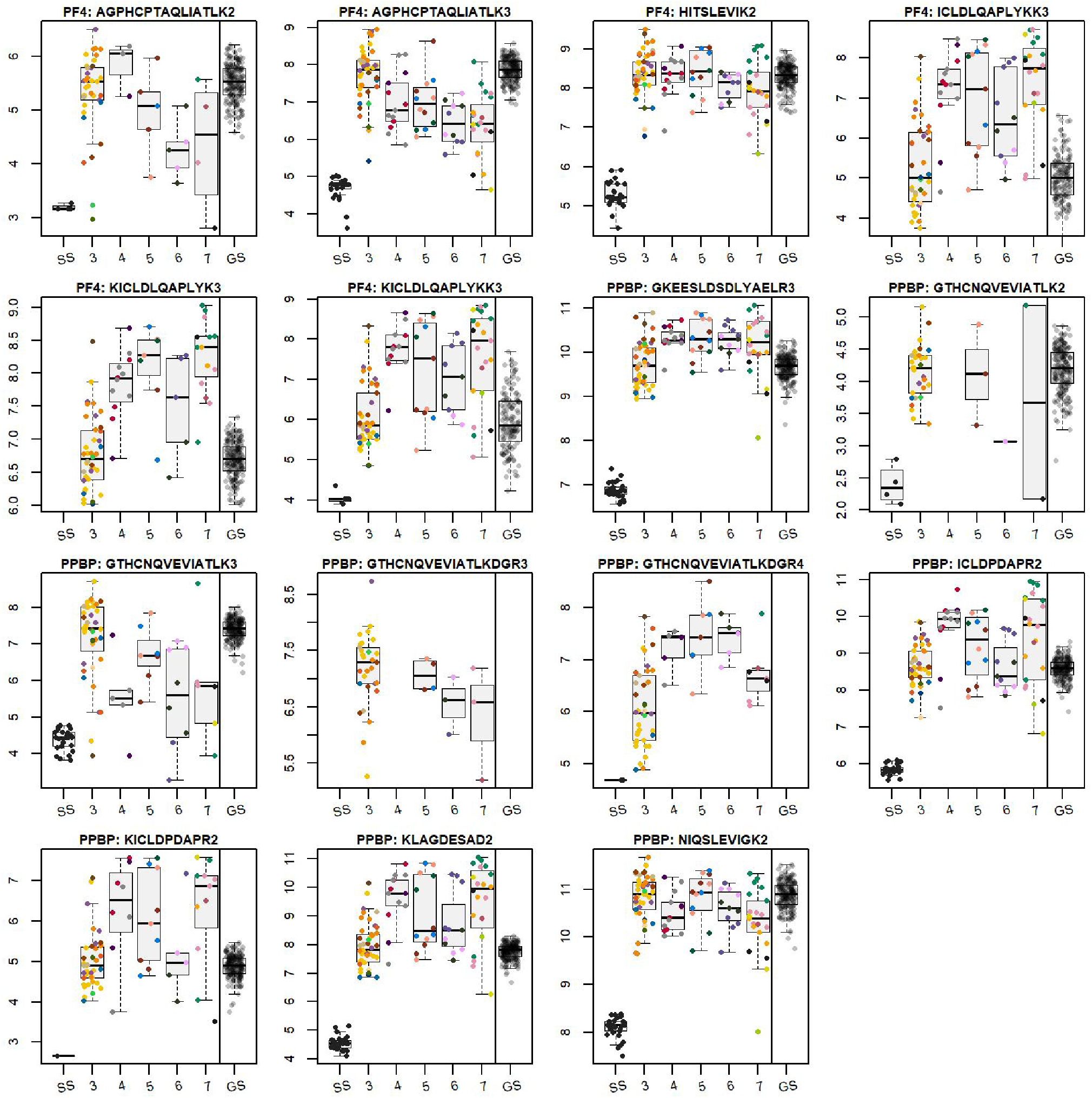
Differential response of peptide precursors mapping to PF4 and PPBP. Boxplots are presented for log_2_-transformed peptide precursor quantities. The peptide precursors charge is indicated with a number after the amino acid sequence. All cysteines were considered carbamidomethylated. Boxplots are drawn based on individual measurements (multiple timepoints per patient), which are indicated with points (different colours for different patients, ordered from left to right by the time taken). Standard Serum sample preparation controls are indicated with “SS” and represent the technical variability (sample preparation + LC-MS). The last box represents the variability seen in the GS study; as the absolute quantities from the two studies cannot be compared directly (the data was acquired on different LC-MS setups), to simplify the visual assessment of the variability, the median of GS quantities was matched to the median of WHO grade 3 (no oxygen support) COVID-19 cases.

**Figure S3.**
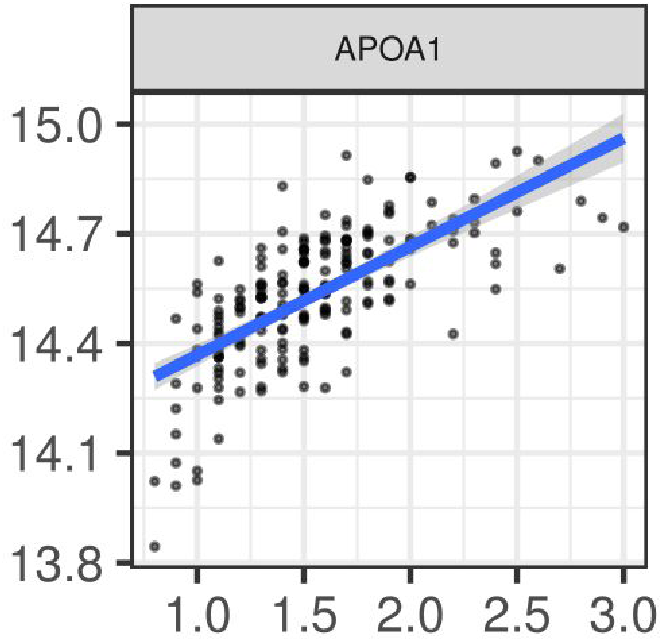
Log_2_-transformed serum levels of Apolipoprotein A1 (APOA1; arbitrary units) plotted against the HDL-Cholesterol concentration (mmol/L) in the GS study.

Figure S4. **Serum protein levels depending on COVID-19 severity (complete list)**. Differentially expressed proteins are highlighted with colours depending on the multiple testing-corrected p-values: green: P < 0.05, yellow: P < 0.01, orange: P < 0.001. In the preprint a PDF will be embedded. Separate .pdf file.

Figure S5. **Age-related protein changes in the general population (GS cohort)**. Separate pdf file.

